# Association of LGBTQ+ Affirming Care With Chronic Disease and Preventive Care Outcomes

**DOI:** 10.1101/2022.05.26.22275633

**Authors:** Tara McKay, Nathaniel M. Tran, Harry Barbee, Judy K. Min

**Author notes:** Address correspondence to: Tara McKay, PhD, Department of Medicine, Health, and Society, Vanderbilt University, 2301 Vanderbilt Pl., PMB 351665, Nashville TN 37235-1665.

## Abstract

**Introduction:** Experiences of discrimination and bias in health care contribute to health disparities for LGBTQ+ and other minority populations. To avoid discrimination, many LGBTQ+ people go to great lengths to find healthcare providers who they trust and are knowledgeable about their health needs. This study examines whether access to an LGBTQ+ affirming provider may improve health outcomes for LGBTQ+ populations across a range of preventive health and chronic disease management outcomes.

**Methods:** This cross-sectional study uses Poisson regression models to examine original survey data (n=1,120) from Wave 1 of the Vanderbilt University Social Networks, Aging, and Policy Study (VUSNAPS), a panel study examining older (50□76 years) LGBTQ+ adults’ health and aging, collected between April 2020 and September 2021.

**Results:** Overall, access to an LGBTQ+ affirming provider is associated with greater uptake of preventive health screenings and improved management of mental health conditions among older LGBTQ+ adults. Compared to participants reporting a usual source of care that is not affirming, participants with an LGBTQ+ affirming provider are more likely to have ever and recently received several types of preventive care, including past year provider visit, flu shot, colorectal cancer screening, and HIV test. Access to an LGBTQ+ affirming provider is also associated with better management of mental health conditions.

**Conclusions:** Inclusive care is essential for reducing health disparities among LGBTQ+ populations. Health systems can reduce disparities by expanding education opportunities for providers regarding LGBTQ+ medicine, adopting nondiscrimination policies for LGBTQ+ patients and employees, and ensuring LGBTQ+ care is included in health insurance coverage.

## INTRODUCTION

Up to one-third of lesbian, gay, bisexual, transgender, and queer (LGBTQ+) Americans avoid seeing a doctor for fear of discrimination.^1,2^ Others go to great lengths to find healthcare providers who they trust, will affirm their identities, and are knowledgeable about their health needs.^3–6^ This study examines the association of access to LGBTQ+ affirming health care with lifetime and timely receipt of preventive health screenings and chronic diseases management outcomes among older LGBTQ+ adults.

While there are no uniform standards for assessing what constitutes LGBTQ+ affirming clinical care, previous work suggests that there are 2 key domains: (1) *clinical competence*, such as understanding specific health needs among LGBTQ+ individuals, and (2) *cultural competence*, which includes respectful communication and interaction with LGBTQ+ individuals.^7–11^ Researchers operationalize LGBTQ+ affirming care at multiple levels (Figure 1), including at the interpersonal level (e.g., during the patient encounter), the institutional level (e.g., the presence of nondiscrimination policies), and the structural level (e.g., laws or rules regulating the provision of gender affirming care). Most research focuses on interpersonal barriers to receiving affirming care, such as disclosure of sexual orientation or gender identity to providers.^12–19^ LGBTQ patients may also experience barriers to accessing affirming care, including lack of insurance or distance to an LGBTQ+ affirming provider.^5,6^

**Figure 1.**
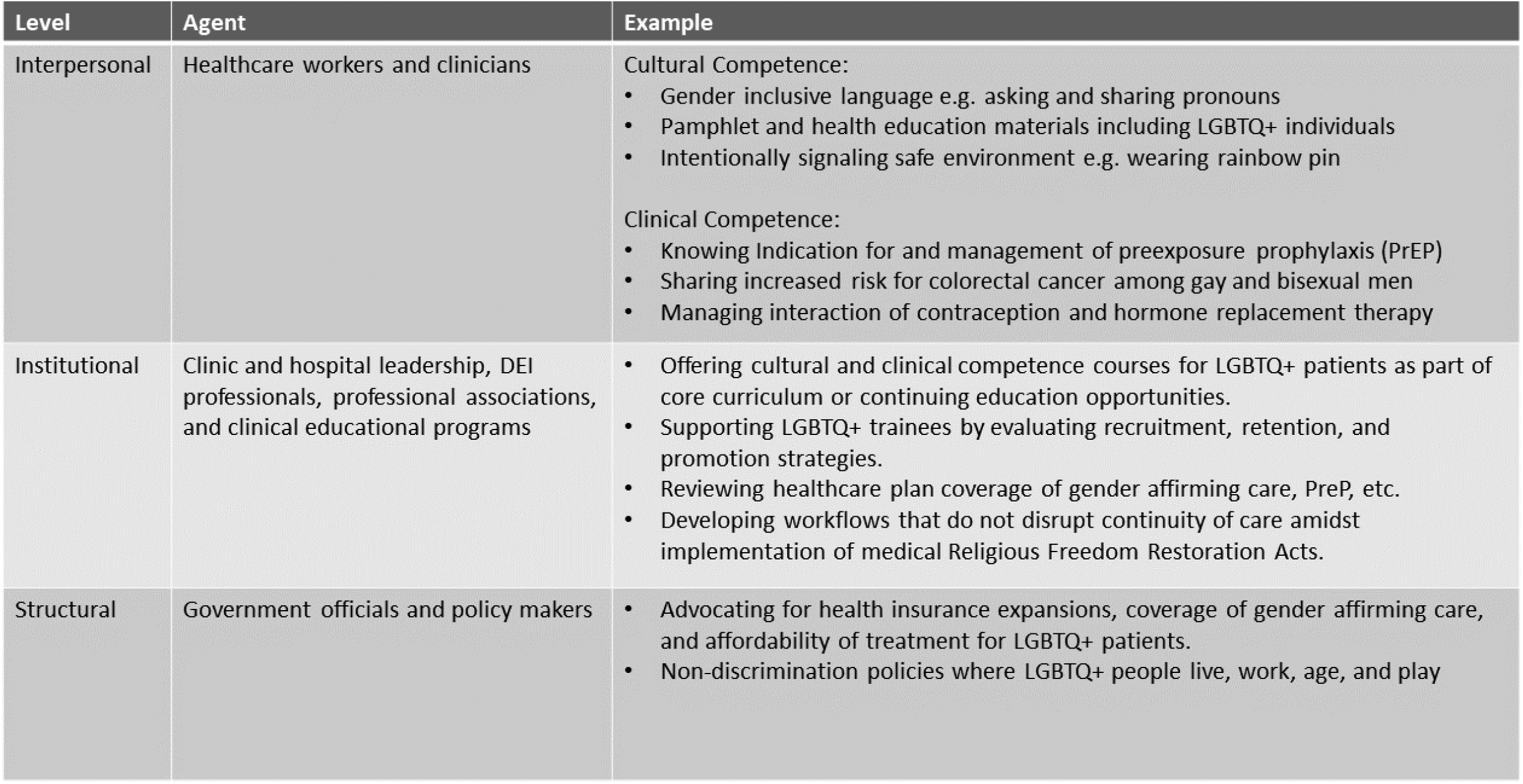
Theoretical model of LGBTQ+ affirming care. LGBTQ+, lesbian, gay, bisexual, transgender, and queer; DEI, diversity, equity, and inclusion.

There are several potential benefits of receiving care from an LGBTQ+ affirming provider. Broadly, affirming care environments may improve health outcomes for LGBTQ+ patients by promoting engagement and retention in care, more timely preventive screenings, provider trust, uptake of provider recommendations, earlier diagnoses, and more open conversations about needs and concerns. As evidence of this, HIV-negative gay and bisexual men with LGBTQ+ affirming providers are more likely to have ever tested for HIV and to be aware of current HIV prevention strategies such as *Undetectable = Untransmitable*.^20^ At the institutional level, patients report higher satisfaction when health systems are LGBTQ+ affirming regardless of sexual orientation or gender identity.^21^ Affirming care practices and explicit nondiscrimination policies may also allow for greater involvement of partners and caregivers in contexts like cancer care, where patients have concerns about disclosing their LGBTQ identity and bringing their partner to appointments.^22–24^

Conversely, when patients do not feel comfortable disclosing or are not asked^19^ about their sexual orientation, gender identity, or sexual behavior, this can lead to inattention to health needs and missed diagnostic screenings.^17,18,25–27^ Gay and bisexual men who do not disclose their sexual orientation to their primary provider are less likely to receive HIV and other STI tests and hepatitis vaccinations.^12,28–30^ The lack of affirming care options for sexual minorities and transgender people can also lead to healthcare fragmentation, where individuals seek care outside of primary care contexts because of gaps in provider knowledge, greater comfort with community providers, or expectations of discrimination.^31,32^ Finally, experiences of disrespect toward a patient’s partner are not uncommon in healthcare settings^33^ and may decrease partner involvement in care decisions.^22–24,34^

This study focuses on older LGBTQ+ adults aged 50 to 76 years, a cohort that has experienced substantial individual, institutional, and structural discrimination throughout their lives.

Consistent with accumulated exposure to minority stressors across the life course, health disparities by sexual orientation and gender identity are especially pronounced at older ages.^35–39^ Experiences of discrimination, stigma, and harassment within healthcare settings reinforce and exacerbate health disparities.^27,40^ By focusing on older adults aged 50 to 76 years, a group that is understudied despite high rates of unmet medical needs,^41^ this study is able to assess both lifetime and timely uptake of many preventive cancer screenings that only are recommended later in life.

Additionally, by focusing on the U.S. South, where an estimated 35% of LGBTQ+ adults in the U.S. live, this study offers insight into how expanding access to LGBTQ+ affirming care might intervene in the poorer health trajectories of LGBTQ people in Southern states,^42^ even as many of these states pursue criminalization of gender affirming care and make allowances for providers to deny care on the basis of religious belief to LGBTQ+ patients.^43^

## METHODS

### Study Sample

This study uses survey data from Wave 1 of the Vanderbilt University Social Networks, Aging, and Policy Study (VUSNAPS; n=1,256), a panel study examining older LGBTQ+ adults’ health and aging, collected between April 2020 and September 2021. Respondents included LGBTQ+ adults aged 50 to 76 years residing in Alabama, Georgia, North Carolina, and Tennessee recruited via community outreach at LGBTQ+ and senior organizations, events, and paid targeted online ads on social media platforms. These states were selected because they reflect the diversity of population and policy relevant to LGBTQ+ populations in the U.S. South. The VUSNAPS Wave 1 sample generally reflects the demographic characteristics of the LGBTQ population aged 50 to 76 years age for sample states and the U.S. South as measured by the U.S. Census Household Pulse Survey (HPS), Phase 3.2, weeks 34_J39 (Appendix Table 1).^44^ Compared to weighted HPS estimates of demographic characteristics of LGBTQ people in the U.S. South, VUSNAPS participants are more educated (71% college degree or higher vs 32% in HPS), less likely to identify as bisexual (11% vs 26%), and less likely to identify as Latino/Hispanic (1% vs 17%). This study was approved by the Vanderbilt University IRB.

Analyses were restricted to LGBTQ+ individuals with a usual source of care to avoid comparing LGBTQ+ affirming care to those without care and to estimate the marginal benefit of having an LGBTQ+ affirming provider versus standard care. The analytic sample included 1,120 respondents, excluding 105 participants who reported the emergency room as their usual source of care and 31 respondents who were missing covariate information on education and/or household income.

### Measures

Respondents were asked “Do you have an LGBT-affirming health care provider?” with response options: “Yes, they are my primary health care provider; Yes, I see them in addition to another health care provider; No, I don’t need or want an LGBT-affirming health care provider; No, I cannot find an LGBT-affirming health care provider in my area; I don’t know; and No answer.” Respondents who reported “Yes” were coded as having access to an LGBTQ+ affirming health care provider. All others were coded as no.

Preventive health outcomes included receipt of timely and lifetime preventive care. Respondents were asked if they had “seen a doctor or health care provider” in the past year. Receipt of lifetime preventive care was assessed by asking, “Have you <u>ever</u> had any of the following preventive care screenings or tests?” including flu shot, breast cancer screening or mammogram (participants assigned female at birth [AFAB]), Pap test (participants AFAB), colorectal cancer screening or colonoscopy, and HIV test (participants who are HIV negative and identify as cisgender male, transgender male, transgender female, or gender non-conforming). If respondents indicated ever having 1 or more of these tests, timely receipt was assessed by asking, “Have you had any of the following tests or screenings <u>in the last 3 years</u>?”). Although screening recommendations vary, the U.S. Centers for Disease Control and Prevention and the U.S. Preventive Services Task Force recommend mammogram screening, cervical cancer screening, and HIV testing at least every 3 years for most adults in this sample.^45^ Colorectal cancer screening is recommended for all adults beginning at age 50 years, and then every 5 to 10 years depending on screening mode and risk factors. Given the age range of the sample, only lifetime receipt of colorectal cancer screening is examined.

Chronic disease management was based on measurement of 5 common conditions with elevated prevalence among LGBTQ+ populations: high blood pressure, diabetes, any heart condition, any respiratory condition, any mental health condition.^46,47^ Conditional on having a specific health condition, respondents were asked: “Is your condition [high blood pressure, diabetes, heart condition, respiratory condition, mental health condition] pretty much under control (1) or is it still a problem (0)?”).

### Statistical Analysis

All analyses were conducted using Stata v17. Chi-square and unadjusted Poisson regression were used to test bivariate differences in the distributions of responses. For binary outcome variables, adjusted prevalence ratios using modified Poisson models with robust error variance were estimated.^48,49^ Adjusted analyses controlled for factors identified in prior research predicting LGBTQ healthcare use and access,^11^ including: age, gender identity, sexual orientation, race/ethnicity, education, income, health insurance status, state of residence, whether the respondent had seen a doctor or healthcare provider in the past year (except where outcome), and the presence of any chronic condition (preventive outcomes only).

## RESULTS

About two-thirds (63%) of respondents had an LGBTQ+ affirming provider. Among the remaining one-third of respondents (N=412), a majority (64.3%) indicated that they did not know if their provider was affirming, 17.5% indicated that they did not need or want and LGBTQ+ affirming provider, and 13.1% could not find one. All respondents reported a usual source of care despite not having an LGBTQ+ affirming provider.

Table 1 presents demographic characteristics of the sample by access to an affirming provider. There were significant differences in the distribution of access to an LGBTQ+ affirming care provider across several demographic characteristics. Most notably, respondents with an LGBTQ+ affirming provider were more likely to: be cisgender men (*p*<0.05) or transgender women (*p*<0.05) than cisgender women; be gay than bisexual (*p*=0.001); have completed a graduate or professional degree than have less than a college degree (*p*=0.05); be living in North Carolina compared with Alabama (*p*=0.05); and be living with HIV (*p*<0.001).

**Table 1.** Sample Demographic Characteristics by Provider LGBTQ+ Affirming Status

|  |  | By access to an LGBTQ+ affirming provider |  |  |
| --- | --- | --- | --- | --- |
|  | Overall | No, non-LGBTQ+ affirming | Yes, LGBTQ+ affirming |  |
| Demographic characteristic | Number (column %) | Number (row %) | Number (row %) | *p*-value |
| Gender identity |  |  |  | **<0.001** |
| Cisgender man | 617 (55.1) | 210 (34.0) | 407 (66.0) |  |
| Cisgender woman | 424 (37.9) | 186 (43.9) | 238 (56.1) |  |
| Transgender woman<sup>a</sup> | 31 (2.8) | 4 (12.9) | 27 (87.1) |  |
| Transgender man<sup>b</sup> | 24 (2.1) | 4 (16.7) | 20 (83.3) |  |
| Transgender/Nonbinary/Gender nonconforming<sup>c</sup> | 24 (2.1) | 8 (33.3) | 16 (66.7) |  |
| Sexual orientation* |  |  |  | **<0.001** |
| Lesbian or gay | 968 (86.4) | 336 (34.7) | 632 (65.3) |  |
| Bisexual | 119 (10.6) | 60 (50.4) | 59 (49.6) |  |
| Not lesbian, gay, or bisexual | 33 (2.9) | 16 (48.5) | 17 (51.5) |  |
| Race and ethnicity |  |  |  | ns |
| White | 973 (86.9) | 345 (35.5) | 628 (64.5) |  |
| Black | 83 (7.4) | 38 (45.8) | 45 (54.2) |  |
| Asian, Hispanic, Multiracial, or Other | 64 (5.7) | 29 (45.3) | 35 (54.7) |  |
| Education |  |  |  | **<0.001** |
| Less than college | 310 (27.7) | 139 (44.8) | 171 (55.2) |  |
| College degree | 353 (31.5) | 137 (38.8) | 216 (61.2) |  |
| Grad/Professional degree | 457 (40.8) | 136 (29.8) | 321 (70.2) |  |
| Household income |  |  |  | **<0.001** |
| <\$45,000 | 292 (26.1) | 128 (43.8) | 164 (56.2) | |
| \$45,000–\$74,999 | 263 (23.5) | 105 (39.9) | 158 (60.1) | |
| \$75,000–\$12,999 | 302 (27.0) | 114 (37.7) | 188 (62.3) | |
| >\$125,000 | 263 (23.5) | 65 (24.7) | 198 (75.3) | |
| State of residency |  |  |  | **<0.01** |
| Alabama | 202 (18.0) | 94 (46.5) | 108 (53.5) |  |
| North Carolina | 322 (28.7) | 97 (30.1) | 225 (69.9) |  |
| Tennessee | 351 (31.3) | 129 (36.8) | 222 (63.2) |  |
| Georgia | 245 (21.9) | 92 (37.6) | 153 (62.4) |  |
| Health insurance |  |  |  | **<0.05** |
| No | 45 (4.0) | 24 (53.3) | 21 (46.7) |  |
| Yes | 1,075.00 (96.0) | 388 (36.1) | 687 (63.9) |  |
| HIV status |  |  |  | **<0.001** |
| Negative or don't know | 987 (88.1) | 396 (40.1) | 591 (59.9) |  |
| Positive | 133 (11.9) | 16 (12.0) | 117 (88.0) |  |
| Any chronic disease |  |  |  | **<0.05** |
| None | 140 (12.5) | 62 (44.3) | 78 (55.7) |  |
| 1 or more | 980 (87.5) | 350 (35.7) | 630 (64.3) |  |
| Total | 1,120.00 (100.0) | 412 (100.0) | 708 (100.0) |  |

Note: Boldface indicates statistical significance. \* $p < 0.1$ , \*\* $p < 0.05$ , \*\*\* $p < 0.01$ LGBTQ+, lesbian, gay, bisexual, transgender, and queer; ns, nonsignificant.
<sup>a</sup>Gender identity was assessed using multiple domains, including current gender identity (multiple selection allowed), sex assigned at birth, a direct “are you transgender?” item, and, for transgender and other gender diverse respondents, daily presentation (man, woman, both, neither). Respondents are coded as transgender women when they identify as transgender and as a woman and were assigned male at birth.
<sup>b</sup>Respondents are coded as transgender men when they identify as transgender and as a man and were assigned female at birth.
<sup>c</sup>Respondents are coded as transgender nonbinary/gender nonconforming when they identify as transgender or nonbinary or gender nonconforming and do not also identify as a cisgender man or cisgender woman.

Table 2 presents adjusted prevalence ratios for the associations among having an LGBTQ+ affirming care provider and preventive care and chronic disease management outcomes. Compared to respondents receiving standard care, respondents with an LGBTQ+ affirming provider were more likely to have ever and recently received several types of preventive care. Individuals with an LGBTQ+ affirming provider were 3.8% (95% CI=1.1%, 6.7%, *p*<0.01) more likely to have seen a doctor in the past year, 7.0% (95% CI=0.1%, 14.3%, *p*<0.05) more likely to have ever had a colorectal cancer screening, 6.0% (95% CI=1.2%, 11.1%, *p*<0.01) more likely to have ever had a flu shot, and, 7.8% (95% CI=2.2%, 13.7%, *p*<0.01) more likely to have had a flu shot in the last 3 years. Among those most at risk of HIV, respondents with an affirming provider were 13.9% (95% CI=3.8%, 24.9%, *p*<0.01) more likely to have ever had an HIV test, and 32.7% (95% CI=7.2%, 64.4%, *p*<0.01) more likely to have an HIV test in the last 3 years. There were no differences in timely or lifetime receipt of Pap test and mammogram screenings among participants assigned female at birth. Figure 2 plots adjusted prevalence ratios for all preventive care and chronic disease management outcomes.

**Table 2.** Unadjusted Prevalence of Preventive Care and Chronic Disease Management Outcomes by Provider LGBTQ+ Affirming Status

|  | Overall | Access to an LGBTQ+ affirming care provider |  | APR (95% CI) | Sample size |
| --- | --- | --- | --- | --- | --- |
|  |  | No, non-LGBTQ+ affirming | Yes, LGBTQ+ affirming |  |  |
| Outcomes of interest | Number (%) | Number (%) | Number (%) |  |  |
| Preventive care |  |  |  |  |  |
| Seen provider, past year***<sup>a</sup> | 1,081 (96.5) | 386 (93.7) | 695 (98.2) | **1.038** (1.011, 1.067) | 1,120 |
| Flu shot, lifetime**<sup>b</sup> | 999 (89.2) | 347 (84.2) | 652 (92.1) | **1.060** (1.012, 1.111) | 1,120 |
| Flu shot, timely*** | 960 (85.7) | 328 (79.6) | 632 (89.3) | **1.078** (1.022, 1.137) | 1,120 |
| Colorectal, lifetime** | 884 (78.9) | 304 (73.8) | 580 (81.9) | **1.070** (1.001, 1.143) | 1,120 |
| HIV test, lifetime***<sup>c</sup> | 440 (78.2) | 154 (73.3) | 286 (81) | **1.139** (1.038, 1.249) | 563 |
| HIV test, timely*** | 245 (43.5) | 80 (38.1) | 165 (46.7) | **1.327** (1.072, 1.644) | 563 |
| Mammogram, lifetime<sup>d</sup> | 428 (93.2) | 180 (93.3) | 248 (93.2) | 0.994 (0.941, 1.051) | 459 |
| Mammogram, timely | 374 (81.5) | 153 (79.3) | 221 (83.1) | 1.034 (0.946, 1.130) | 459 |
| Pap test, lifetime | 400 (87.1) | 171 (88.6) | 229 (86.1) | 0.971 (0.903, 1.044) | 459 |
| Pap test, timely | 269 (58.6) | 118 (61.1) | 151 (56.8) | 0.912 (0.777, 1.070) | 459 |
| Chronic disease management |  |  |  |  |  |
| Mental health condition* | 386 (73) | 126 (65.3) | 260 (77.4) | **1.108** (0.986, 1.244) | 529 |
| Blood pressure | 550 (91.8) | 203 (89.4) | 347 (93.3) | 1.035 (0.979, 1.095) | 599 |
| Diabetes | 175 (73.8) | 72 (75) | 103 (73) | 0.979 (0.826, 1.159) | 237 |
| Heart condition | 144 (88.3) | 65 (90.3) | 79 (86.8) | 1.019 (0.902, 1.152) | 163 |
| Respiratory condition | 211 (86.1) | 87 (82.9) | 124 (88.6) | 0.998 (0.896, 1.112) | 245 |
| Arthritis or rheumatism | 169 (50) | 69 (50) | 100 (50) | 0.957 (0.759, 1.207) | 338 |

Note: Data come from Wave I VUSNAPS (R01AG063771). Boldface indicates statistical significance. \* $p < 0.1$ , \*\* $p < 0.05$ , \*\*\* $p < 0.01$ . LGBTQ+, lesbian, gay, bisexual, transgender, and queer; ns, nonsignificant. Number is the raw count and % is the raw proportion. APR is the adjusted Prevalence Ratio adjusting for gender identity, race and ethnicity, sexual orientation, age, household income, education, state of residency and insurance status.
<sup>a</sup>All preventive care outcomes with the exception of HIV testing outcomes were adjusted for having any chronic condition, including high blood pressure or hypertension, diabetes or high blood sugar, heart attack or coronary heart disease, asthma, arthritis or rheumatism, a mental health condition, cancer, stroke, or HIV.
<sup>b</sup>All preventive and chronic disease management outcomes besides seeing a doctor in the last year were adjusted for having seen a doctor in the last year.
<sup>c</sup>HIV testing outcomes were restricted to cisgender men, transgender men, transgender women, and gender non- conforming individuals.
<sup>d</sup>Mammogram and Pap test outcomes were restricted to participants assigned female at birth.

**Figure 2.**
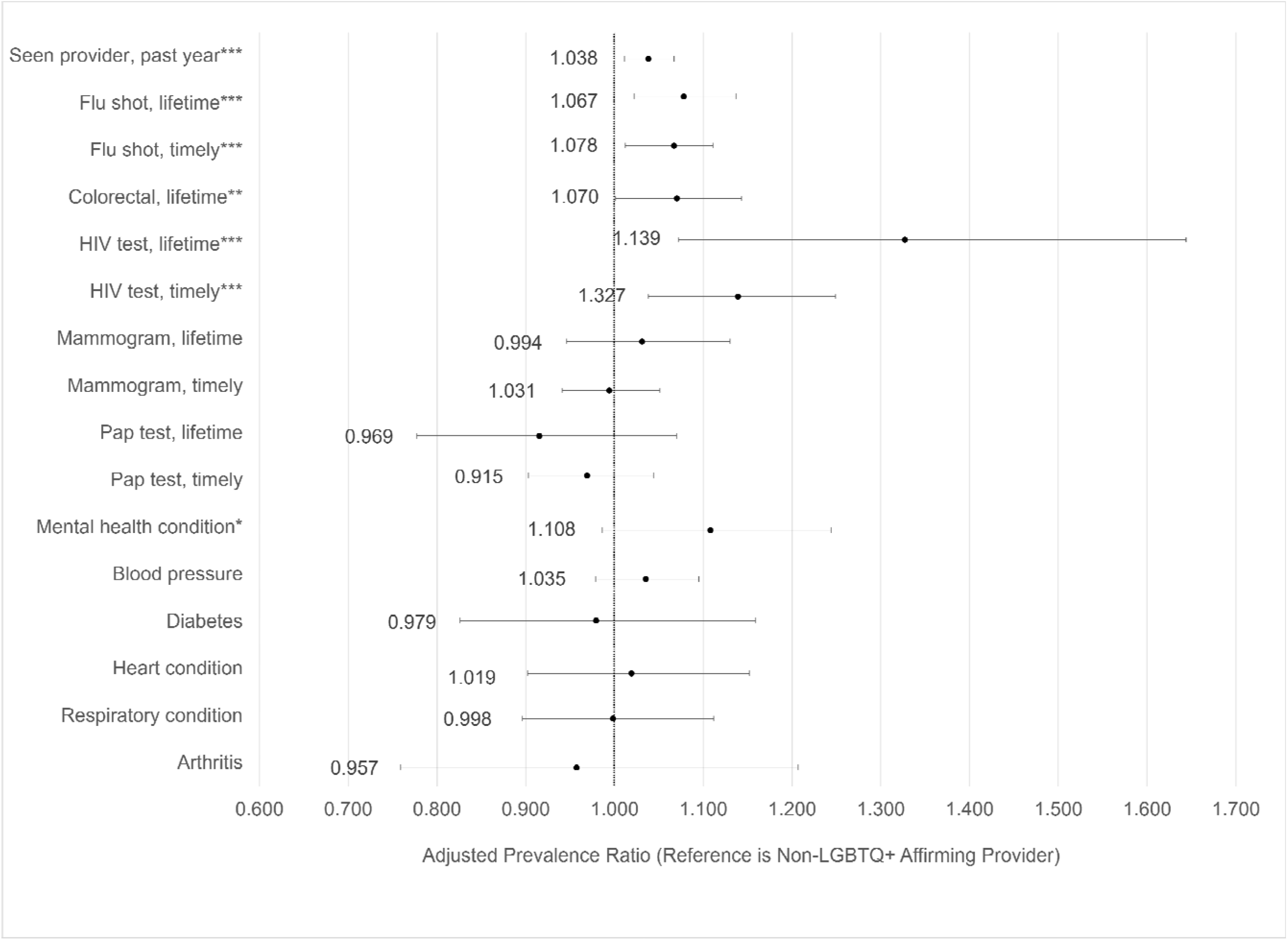
Adjusted prevalence ratios of preventive care and chronic disease management outcomes by access to LGBTQ+ affirming provider. Note: * p<.10 **p<.05 ***p<.01. LGBTQ+, lesbian, gay, bisexual, transgender, and queer. Source: Vanderbilt University Social Network, Aging, and Policy Study (VUSNAPS), Wave I (N=1,120).

LGBTQ+ individuals with a mental health condition were 10.8% (95% CI= □1.4%, 24.4%, *p*<0.10) more likely to report their condition “under control” when they had an affirming provider versus standard care. There were no significant differences by provider type in the likelihood that respondents reported other health conditions as “under control.”

## DISCUSSION

Access to an LGBTQ+ affirming provider varied across demographic characteristics and was associated with receipt of several preventive care outcomes and better patient-reported management of mental health conditions. Respondents who had LGBTQ+ affirming healthcare provider were more likely to have had a doctor’s visit in the past year, to have ever had a flu shot, and to have ever had a colorectal cancer screening compared to LGBTQ+ adults receiving standard care.

LGBTQ+ adults with a mental health condition were also more likely to report that their mental health condition was under control when they had an affirming provider. This finding is consistent with other work showing the benefits of accessing gender affirming care for mental health among transgender young people^50–53^ and adults.^54^

Notably, those most at risk of HIV were more likely to have ever had an HIV test and more likely to have had an HIV test in the last 3 years when they had an LGBTQ+ affirming provider. More than half of new HIV infections in the U.S. occur in the South, and people living with HIV are more than 3 times more likely to die from the disease compared to the rest of the country.^56^ Links between affirming care and HIV prevention outcomes in this and other studies^20^ help us better understand the poorer HIV trajectories of the U.S. South, where LGBTQ patients face a dearth of LGBTQ affirming providers^55^ and are more likely to live in states with laws that deny or limit health care for sexual and gender minorities.^43^

Respondents with an affirming care provider also had higher rates of lifetime receipt of colorectal cancer screening, most of which are conducted via colonoscopy.^57^ While there may be several aspects of affirming care that are relevant to this outcome, related work suggests that cultural competency and trust are especially important for acceptability and uptake of invasive procedures like colonoscopy among minority populations.^58^

Although sexual minority women are less likely to be offered a Pap test than heterosexual women,^59^ there were no associations between having an LGBTQ+ affirming provider and lifetime or timely receipt of Pap test or mammogram screenings among LGBTQ+ individuals AFAB. This is likely explained by several factors. The study does not include information on current anatomy; thus, analyses may include individuals for whom these screenings would be inappropriate. Patient care and research with transgender and gender diverse populations should adopt anatomical inventories in electronic health records and study measures to improve accuracy and patient-centeredness of care.^60^ Additionally, Pap tests and mammograms have higher rates of completion compared with other screenings,^57^ longer lifetime risk exposure (Pap test), and higher acceptability compared to HIV tests or colorectal cancer screenings.^61–64^

There are several behaviors that may promote higher uptake of preventive care and better mental health management among individuals with an affirming provider, including the use of visual cues, such as a rainbow pin,^65^ or communicating to cisgender gay and bisexual men that they are at elevated risk for developing colorectal cancer.^66^ LGBTQ+ affirming providers may also support patient uptake of invasive procedures by using gender-neutral language when asking patients whether they have a spouse or partner who can provide transportation and support afterward. By engaging in these varied practices, LGBTQ+ affirming clinicians and institutions may allow for earlier diagnoses, more open conversations about patient concerns, and greater patient uptake of provider recommendations. Additionally, among populations where many are hesitant or lack trust in their healthcare providers,^35,67^ retention in care, measured here as having had a recent visit to a usual source of care, is a major achievement.

Health systems should prioritize LGBTQ+ inclusive best practices to achieve health equity for LGBTQ+ populations. Best practices identified by the American Medical Association include the visible display of information and images that reflect and center LGBTQ identities and health concerns, and the collection of sexual orientation and gender identity on patient intake forms.^68,69^ In addition, institutions should adopt higher level policy statements on nondiscrimination of LGBTQ patients and employees; provide gender-neutral restroom facilities; engage openly with LGBT referral networks and identify LGBT-specific competencies on institution websites; develop protocols to support to LGBTQ physicians, staff, and patients who encounter antagonistic patients in clinical spaces; and expand formal continuing medical education (CME) offerings and staff development trainings on inclusive language use, LGBTQ+ identities, family structures, behaviors, and health needs beyond sexual health.^70–73^ In general, physicians and nurses have few or no reservations about providing care to LGBTQ+ populations; however, they often feel unprepared to support LGBTQ+ patients across a range of health needs (emergency medicine, cancer care, dementia care, palliative care, etc.) given limited engagement with LGBTQ+ health in medical curricula,^74–79^ even when they also identify as LGBT.^33^ These changes will be a first step toward improving LGBTQ+ engagement with preventive services, reducing healthcare fragmentation, and ensuring the opportunity of access to all members of the LGBTQ+ community.

Longer-term, health systems can improve training and retention opportunities for LGBTQ+ health professionals by updating nondiscrimination policies to explicitly include sexual orientation and gender identity, and expanding fellowship and residency opportunities in LGBTQ+ health and medicine. Despite improvements since the 1990s, LGBT physicians continue to experience discrimination in the workplace, are denied referrals from colleagues, and witness discrimination against LGBT employees, patients, and patients’ partners.^33^ Physician workforce diversity matters for patient outcomes and reducing health disparities. Rigorous studies demonstrate that having a gender- or race-match between doctors and patients reduces mortality and adverse outcomes in hospital settings, increases uptake of preventive care, and increases patient satisfaction.^80,58,81,82^ While not all LGBTQ+ patients may need or want to access an LGBTQ+ provider, LGBTQ+ health disparities may be improved by increasing training opportunities in LGBTQ+ medicine and for LGBTQ+ physicians, and by decreasing experiences of discrimination on the job that threaten retention of LGBTQ+ health professionals.^33^

## Limitations

While this study offers critical insight into potential benefits associated with access to an LGBTQ+ affirming provider for older LGBTQ+ adults in the U.S. South, there were some limitations. First, the item asking whether respondents had an LGBTQ+ affirming provider did not include a definition or example. LGBTQ+ adults may have had different perceptions of what “affirming” means in the context of health care. Future work should examine how patients identify whether their provider is culturally and clinically competent in LGBTQ+ health needs.

Second, while the VUSNAPS sample broadly reflects the characteristics of older LGBTQ+ adults in the U.S. South, the sample underrepresents the experiences of racial/ethnic minority, bisexual, and less educated members of the LGBTQ+ community who are often more likely to experience discrimination or erasure across multiple minority statuses, live in geographic locations with fewer affirming providers, and experience higher barriers to accessing care overall.^59,83,84^

Finally, data collection overlapped with the earliest period of the COVID-19 pandemic. The study mitigated the effects of the pandemic by measuring preventive care outcomes in the last 3 years to capture utilization prior to the onset of the pandemic. However, pandemic-related healthcare delays or restrictions may have still disrupted some preventive care.

## CONCLUSIONS

Expanding access to LGBTQ+ affirming providers across the health system may help narrow disparities in morbidity for LGBTQ+ older adults and intervene in the poorer health trajectories of LGBTQ people in many Southern states. To increase access to LGBTQ+ affirming providers, medical education and healthcare systems must expand formal and continuing education opportunities around LGBTQ+ medicine and adopt best practices for LGBTQ+ affirming care.

## Data Availability

Deidentified data used by the present study are available upon reasonable request to the first author. Additional information may be found at www.vusnaps.com.

## ACKNOWLEDGMENTS

We are grateful to members of the Vanderbilt LGBT Policy Lab for their valuable feedback on this work and to VUSNAPS participants. This work was supported by the National Institute on Aging (5R01AG063771-03, 3R01AG063771-03S1), Vanderbilt University, and Vanderbilt University Medical Center.

Funding provided by NIA 5R01AG063771-03 and 3R01AG063771-03S1.

The authors have no financial disclosures.

---

**Appendix Table 1.**
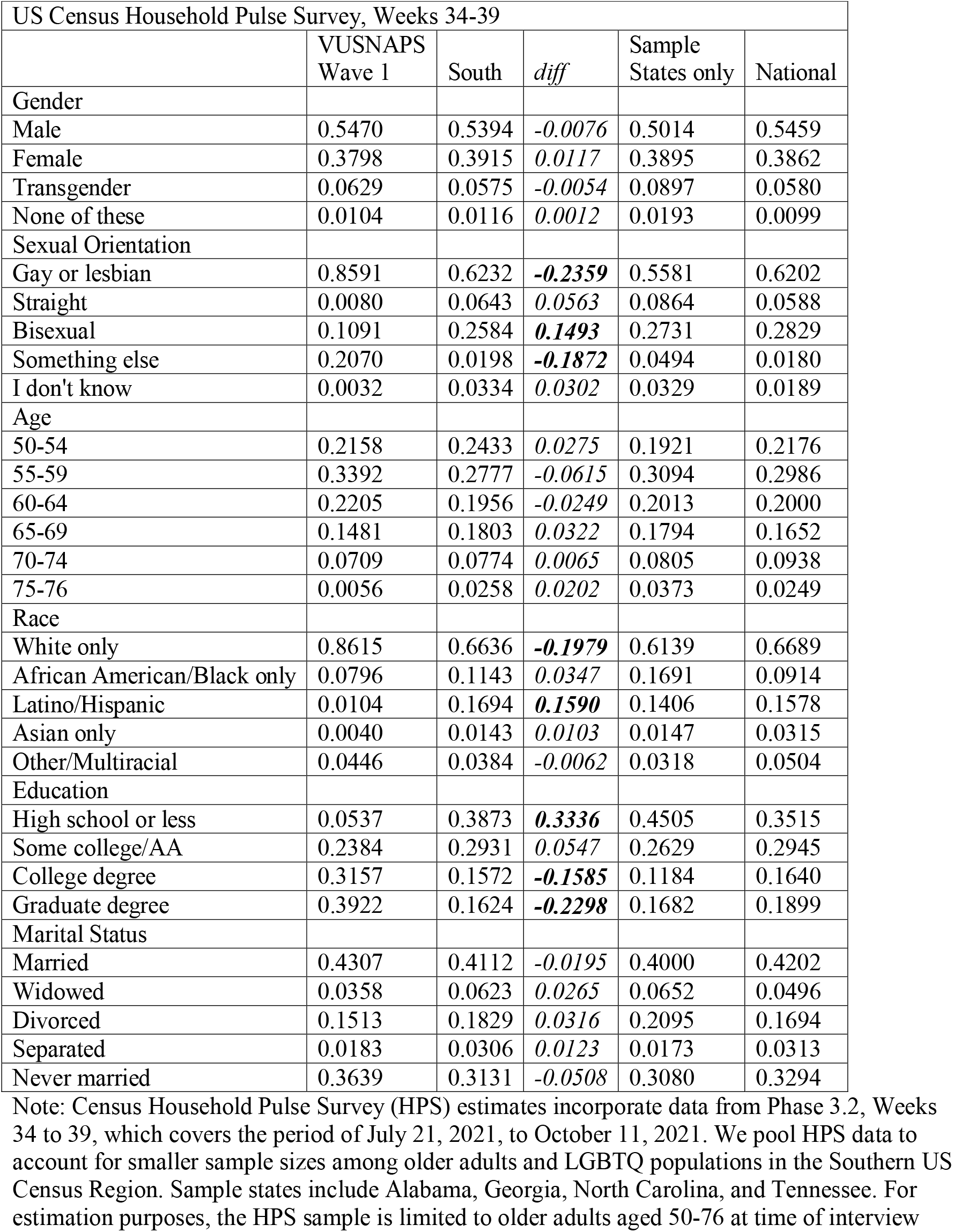
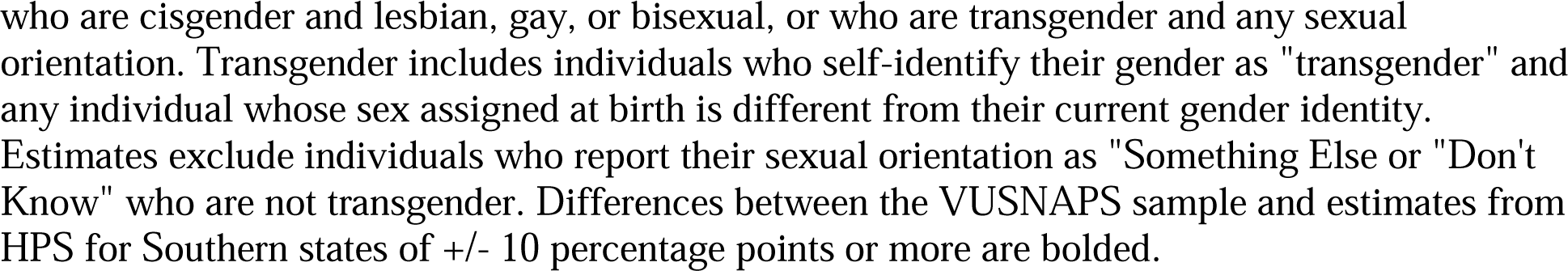
Comparison of VUSNAPS Sample Characteristics with Census Household Pulse Survey Estimates for LGBT Adults Aged 50-76 in 2021.

| US Census Household Pulse Survey, Weeks 34-39 |  |  |  |  |  |
| --- | --- | --- | --- | --- | --- |
|  | VUSNAPS Wave 1 | South | *diff* | Sample States only | National |
| Gender |  |  |  |  |  |
| Male | 0.5470 | 0.5394 | *-0.0076* | 0.5014 | 0.5459 |
| Female | 0.3798 | 0.3915 | *0.0117* | 0.3895 | 0.3862 |
| Transgender | 0.0629 | 0.0575 | *-0.0054* | 0.0897 | 0.0580 |
| None of these | 0.0104 | 0.0116 | *0.0012* | 0.0193 | 0.0099 |
| Sexual Orientation |  |  |  |  |  |
| Gay or lesbian | 0.8591 | 0.6232 | ***-0.2359*** | 0.5581 | 0.6202 |
| Straight | 0.0080 | 0.0643 | *0.0563* | 0.0864 | 0.0588 |
| Bisexual | 0.1091 | 0.2584 | ***0.1493*** | 0.2731 | 0.2829 |
| Something else | 0.2070 | 0.0198 | ***-0.1872*** | 0.0494 | 0.0180 |
| I don't know | 0.0032 | 0.0334 | *0.0302* | 0.0329 | 0.0189 |
| Age |  |  |  |  |  |
| 50-54 | 0.2158 | 0.2433 | *0.0275* | 0.1921 | 0.2176 |
| 55-59 | 0.3392 | 0.2777 | *-0.0615* | 0.3094 | 0.2986 |
| 60-64 | 0.2205 | 0.1956 | *-0.0249* | 0.2013 | 0.2000 |
| 65-69 | 0.1481 | 0.1803 | *0.0322* | 0.1794 | 0.1652 |
| 70-74 | 0.0709 | 0.0774 | *0.0065* | 0.0805 | 0.0938 |
| 75-76 | 0.0056 | 0.0258 | *0.0202* | 0.0373 | 0.0249 |
| Race |  |  |  |  |  |
| White only | 0.8615 | 0.6636 | ***-0.1979*** | 0.6139 | 0.6689 |
| African American/Black only | 0.0796 | 0.1143 | *0.0347* | 0.1691 | 0.0914 |
| Latino/Hispanic | 0.0104 | 0.1694 | ***0.1590*** | 0.1406 | 0.1578 |
| Asian only | 0.0040 | 0.0143 | *0.0103* | 0.0147 | 0.0315 |
| Other/Multiracial | 0.0446 | 0.0384 | *-0.0062* | 0.0318 | 0.0504 |
| Education |  |  |  |  |  |
| High school or less | 0.0537 | 0.3873 | ***0.3336*** | 0.4505 | 0.3515 |
| Some college/AA | 0.2384 | 0.2931 | *0.0547* | 0.2629 | 0.2945 |
| College degree | 0.3157 | 0.1572 | ***-0.1585*** | 0.1184 | 0.1640 |
| Graduate degree | 0.3922 | 0.1624 | ***-0.2298*** | 0.1682 | 0.1899 |
| Marital Status |  |  |  |  |  |
| Married | 0.4307 | 0.4112 | *-0.0195* | 0.4000 | 0.4202 |
| Widowed | 0.0358 | 0.0623 | *0.0265* | 0.0652 | 0.0496 |
| Divorced | 0.1513 | 0.1829 | *0.0316* | 0.2095 | 0.1694 |
| Separated | 0.0183 | 0.0306 | *0.0123* | 0.0173 | 0.0313 |
| Never married | 0.3639 | 0.3131 | *-0.0508* | 0.3080 | 0.3294 |

Note: Census Household Pulse Survey (HPS) estimates incorporate data from Phase 3.2, Weeks 34 to 39, which covers the period of July 21, 2021, to October 11, 2021. We pool HPS data to account for smaller sample sizes among older adults and LGBTQ populations in the Southern US Census Region. Sample states include Alabama, Georgia, North Carolina, and Tennessee. For estimation purposes, the HPS sample is limited to older adults aged 50-76 at time of interview
who are cisgender and lesbian, gay, or bisexual, or who are transgender and any sexual orientation. Transgender includes individuals who self-identify their gender as "transgender" and any individual whose sex assigned at birth is different from their current gender identity. Estimates exclude individuals who report their sexual orientation as "Something Else or "Don't Know" who are not transgender. Differences between the VUSNAPS sample and estimates from HPS for Southern states of +/- 10 percentage points or more are bolded.

## REFERENCES

1. NPR, Robert Wood Johnson Foundation, Harvard T.H. Chan School of Public Health. Discrimination in America: Experiences and Views of LGBTQ Americans. Harvard T.H. Chan School of Public Health; 2017. https://cdn1.sph.harvard.edu/wp-content/uploads/sites/94/2017/11/NPR-RWJF-HSPH-Discrimination-LGBTQ-Final-Report.pdf.

2. Ceres M, Quinn GP, Loscalzo M, Rice D. Cancer Screening Considerations and Cancer Screening Uptake for Lesbian, Gay, Bisexual, and Transgender Persons. Semin Oncol Nurs. 2018;34(1):37–51. https://doi.org/10.1016/j.soncn.2017.12.001.

3. Barbara AM, Quandt SA, Anderson RT. Experiences of Lesbians in the Health Care Environment. Women Health. 2001;34(1):45–62. https://doi.org/10.1300/J013v34n01_04.

4. Martos AJ, Fingerhut A, Wilson PA, Meyer IH. Utilization of LGBT-Specific clinics and providers across three cohorts of lesbian, gay, and bisexual people in the United States. SSM Popul Health. 2019;9:100505. https://doi.org/10.1016/j.ssmph.2019.100505.

5. Tuller D. For LGBTQ Patients, High-Quality Care In A Welcoming Environment. Health Aff (Millwood). 2020;39(5):736–739. https://doi.org/10.1377/hlthaff.2020.00345.

6. Giano Z, Hubach RD, Meyers HJ, et al. Assessing the Health Care Experiences of Rural Men Who Have Sex with Men (MSM). J Health Care Poor Underserved. 2020;31(1):235–248. https://doi.org/10.1353/hpu.2020.0020.

7. Keuroghlian AS, Ard KL, Makadon HJ. Advancing health equity for lesbian, gay, bisexual and transgender (LGBT) people through sexual health education and LGBT-affirming health care environments. Sex Health. 2017;14(1):119–122. https://doi.org/10.1071/SH16145.

8. Obedin-Maliver J, Goldsmith ES, Stewart L. Lesbian, Gay, Bisexual, and Transgender-Related Content in Undergraduate Medical Education. JAMA. 2011;306(9):971–977. https://doi.org/10.1001/jama.2011.1255.

9. Sutter ME, Simmons VN, Sutton SK. Oncologists’ experiences caring for LGBTQ patients with cancer: Qualitative analysis of items on a national survey. Patient Educ Couns. 2021;104(4):871–876. https://doi.org/10.1016/j.pec.2020.09.022.

10. Khalili J, Leung LB, Diamant AL. Finding the Perfect Doctor: Identifying Lesbian, Gay, Bisexual, and Transgender-Competent Physicians. Am J Public Health. 2015;105(6). https://doi.org/10.2105/AJPH.2014.302448.

11. Parameshwaran V, Cockbain BC, Hillyard M, Price J. Is the Lack of Specific Lesbian, Gay, Bisexual, Transgender and Queer/Questioning (LGBTQ) Health Care Education in Medical School a Cause for Concern? Evidence From a Survey of Knowledge and Practice Among UK Medical Students. J Homosex. 2017;64(3):367–381. https://doi.org/10.1080/00918369.2016.1190218.

12. Petroll AE, Mosack KE. Physician awareness of sexual orientation and preventive health recommendations to men who have sex with men. Sex Transm Dis. 2011;38(1):63–67. https://doi.org/10.1097/OLQ.0b013e3181ebd50f.

13. Rossman K, Salamanca P, Macapagal K. A Qualitative Study Examining Young Adults’ Experiences of Disclosure and Nondisclosure of LGBTQ Identity to Health Care Providers. 2017;64(10):1390–1410. https://doi.org/10.1080/00918369.2017.1321379.

14. Meckler GD, Elliott MN, Kanouse DE, Beals KP, Schuster MA. Nondisclosure of Sexual Orientation to a Physician Among a Sample of Gay, Lesbian, and Bisexual Youth. Arch Pediatr Adolesc Med. 2006;160(12):1248–1254. https://doi.org/10.1001/archpedi.160.12.1248.

15. Ye M, Kahana B, Kahana E, Xu S. Trust in the Doctor-Patient Relationship Enhances Well-being and Life Satisfaction among Older LGBT People [abstract only]. Innov Aging. 2018;2(Suppl 1):123. https://doi.org/10.1093/geroni/igy023.451.

16. Stein GL, Bonuck KA. Original Research: Physician-Patient Relationships Among the Lesbian and Gay Community. J Gay Lesbian Med Assoc. 2001;5(3):87–93. https://doi.org/10.1023/A:1011648707507.

17. Durso LE, Meyer IH. Patterns and Predictors of Disclosure of Sexual Orientation to Healthcare Providers among Lesbians, Gay Men, and Bisexuals. Sex Res Soc Policy. 2013;10(1):35–42. https://doi.org/10.1007/s13178-012-0105-2.

18. Kamen CS, Smith-Stoner M, Heckler CE, Flannery M, Margolies L. Social support, self-rated health, and lesbian, gay, bisexual, and transgender identity disclosure to cancer care providers. Oncol Nurs Forum. 2015;42(1):44–51. https://doi.org/10.1188/15.ONF.44-51.

19. Boehmer U, Case P. Physicians don’t ask, sometimes patients tell. Cancer. 2004;101(8):1882–1889. https://doi.org/10.1002/cncr.20563.

20. McKay T, Akré ER, Henne J, Kari N, Conway A, Gothelf I. LGBTQ+ Affirming Care May Increase Awareness and Understanding of Undetectable = Untransmittable among Midlife and Older Gay and Bisexual Men in the US South. Int J Environ Res Public Health. 2022;19(17):10534. https://doi.org/10.3390/ijerph191710534.

21. DiLeo R, Borkowski N, O’Connor SJ, Datti P, Weech-Maldonado R. The Relationship Between “Leader in LGBT Healthcare Equality” Designation and Hospitals’ Patient Experience Scores. J Healthc Manag Am Coll Healthc Exec. 2020;65(5):366–377. https://doi.org/10.1097/JHM-D-19-00177.

22. Katz A. Gay and lesbian patients with cancer. Oncol Nurs Forum. 2009;36(2):203–207. https://doi.org/10.1188/09.ONF.203-207.

23. Katz A. Gay and Lesbian Patients with Cancer. In: Mulhall JP, Incrocci L, Goldstein I, Rosen R, eds. Cancer and Sexual Health. Current Clinical Urology. Humana Press; 2011:397–403. https://doi.org/10.1007/978-1-60761-916-1_26.

24. Cloyes KG, Hull W, Davis A. Palliative and End-of-Life Care for Lesbian, Gay, Bisexual, and Transgender (LGBT) Cancer Patients and Their Caregivers. Semin Oncol Nurs. 2018;34(1):60–71. https://doi.org/10.1016/j.soncn.2017.12.003.

25. Makadon HJ. Improving Health Care for the Lesbian and Gay Communities. N Engl J Med. 2009;354(9):895–897. https://doi.org/10.1056/NEJMp058259.

26. Mayer KH, Bradford JB, Makadon HJ, Stall R, Goldhammer H, Landers S. Sexual and gender minority health: what we know and what needs to be done. Am J Public Health. 2008;98(6):989–995. https://doi.org/10.2105/AJPH.2007.127811.

27. Romanelli M, Lindsey MA. Patterns of Healthcare Discrimination Among Transgender Help-Seekers. Am J Prev Med. 2020;58(4):e123–e131. https://doi.org/10.1016/j.amepre.2019.11.002.

28. Ng BE, Moore D, Michelow W, et al. Relationship between disclosure of same-sex sexual activity to providers, HIV diagnosis and sexual health services for men who have sex with men in Vancouver, Canada. Can J Public Health. 2014;105(3):e186–e191. https://doi.org/10.17269/cjph.105.4212.

29. Stupiansky NW, Liau A, Rosenberger J, et al. Young Men’s Disclosure of Same Sex Behaviors to Healthcare Providers and the Impact on Health: Results from a US National Sample of Young Men Who Have Sex with Men. AIDS Patient Care. 2017;31(8):342–347. https://doi.org/10.1089/apc.2017.0011.

30. Metheny N, Stephenson R. Disclosure of Sexual Orientation and Uptake of HIV Testing and Hepatitis Vaccination for Rural Men Who Have Sex With Men. Ann Fam Med. 2016;14(2):155–158. https://doi.org/10.1370/afm.1907.

31. Griffin M, Krause KD, Kapadia F, Halkitis PN. A Qualitative Investigation of Healthcare Engagement Among Young Adult Gay Men in New York City: A P18 Cohort Substudy. LGBT Health. 2018;5(6):368–374. https://doi.org/10.1089/lgbt.2017.0015.

32. Koehler A, Strauss B, Briken P, Szuecs D, Nieder TO. Centralized and Decentralized Delivery of Transgender Health Care Services: A Systematic Review and a Global Expert Survey in 39 Countries. Front Endocrinol. 2021;12:717914. https://doi.org/10.3389/fendo.2021.717914.

33. Eliason MJ, Dibble SL, Robertson PA. Lesbian, gay, bisexual, and transgender (LGBT) physicians’ experiences in the workplace. J Homosex. 2011;58(10):1355–1371. https://doi.org/10.1080/00918369.2011.614902.

34. Seay J, Mitteldorf D, Yankie A, Pirl WF, Kobetz E, Schlumbrecht M. Survivorship care needs among LGBT cancer survivors. J Psychosoc Oncol. 2018;36(4):393–405. https://doi.org/10.1080/07347332.2018.1447528.

35. Fredriksen-Goldsen KI, Kim HJ, Barkan SE, Muraco A, Hoy-Ellis CP. Health disparities among lesbian, gay, and bisexual older adults: results from a population-based study. Am J Public Health. 2013;103(10):1802–1809. https://doi.org/10.2105/AJPH.2012.301110.

36. Fredriksen-Goldsen KI, Kim HJ, Bryan AEB, Shiu C, Emlet CA. The Cascading Effects of Marginalization and Pathways of Resilience in Attaining Good Health Among LGBT Older Adults. Gerontologist. 2017;57(suppl 1):S72–S83. https://doi.org/10.1093/geront/gnw170.

37. Gonzales G, Henning-Smith C. Disparities in health and disability among older adults in same-sex cohabiting relationships. J Aging Health. 2015;27(3):432–453. https://doi.org/10.1177/0898264314551332.

38. Fredriksen-Goldsen KI, Cook-Daniels L, Kim HJ, et al. Physical and Mental Health of Transgender Older Adults: An At-Risk and Underserved Population. Gerontologist. 2014;54(3):488–500. https://doi.org/10.1093/geront/gnt021.

39. Correro AN, Nielson KA. A review of minority stress as a risk factor for cognitive decline in lesbian, gay, bisexual, and transgender (LGBT) elders. J Gay Lesbian Ment Health. 2019;24(1):2–19. https://doi.org/10.1080/19359705.2019.1644570.

40. Reisner SL, Pardo ST, Gamarel KE, Hughto JMW, Pardee DJ, Keo-Meier CL. Substance use to cope with stigma in healthcare among U.S. female-to-male trans masculine adults. LGBT Health. 2015;2(4):324–332. https://doi.org/10.1089/lgbt.2015.0001.

41. IOM. The Health of Lesbian, Gay, Bisexual, and Transgender People: Building a Foundation for a Better Understanding. Published online 2011.

42. Hasenbush A, Flores AR, Kastanis A, Sears B, Gates GJ. The LGBT Divide: A Data Portrait of LGBT People in the Midwestern, Mountain & Southern States. Published online 2014.

43. Movement Advancement Project. Equality Maps: Healthcare Laws and Policies. Published 2021. https://www.lgbtmap.org/equality-maps/healthcare_laws_and_policies.

44. US Census Bureau. Household Pulse Survey Data Tables. https://Census.gov. Accessed July 7, 2022. https://www.census.gov/programs-surveys/household-pulse-survey/data.html.

45. US Centers for Disease Control and Prevention. A and B Recommendations | United States Preventive Services Taskforce. Accessed March 24, 2022. https://www.uspreventiveservicestaskforce.org/uspstf/recommendation-topics/uspstf-and-b-recommendations.

46. Hughes L, Shireman TI, Hughto J. Privately Insured Transgender People Are At Elevated Risk For Chronic Conditions Compared With Cisgender Counterparts. Health Aff (Millwood). 2021;40(9):1440–1448. https://doi.org/10.1377/hlthaff.2021.00546.

47. Fredriksen-Goldsen KI, Kim HJ, Shui C, Bryan AEB. Chronic Health Conditions and Key Health Indicators Among Lesbian, Gay, and Bisexual Older US Adults, 2013-2014. Am J Public Health. 2017;107(8):1332–1338. https://doi.org/10.2105/AJPH.2017.303922.

48. McNutt LA, Wu C, Xue X, Hafner JP. Estimating the Relative Risk in Cohort Studies and Clinical Trials of Common Outcomes. Am J Epidemiol. 2003;157(10):940–943. https://doi.org/10.1093/aje/kwg074.

49. Zou G. A Modified Poisson Regression Approach to Prospective Studies with Binary Data. Am J Epidemiol. 2004;159(7):702–706. https://doi.org/10.1093/aje/kwh090.

50. Tordoff DM, Wanta JW, Collin A, Stepney C, Inwards-Breland DJ, Ahrens K. Mental Health Outcomes in Transgender and Nonbinary Youths Receiving Gender-Affirming Care. JAMA Netw Open. 2022;5(2):e220978. https://doi.org/10.1001/jamanetworkopen.2022.0978.

51. Turban JL, King D, Carswell JM, Keuroghlian AS. Pubertal Suppression for Transgender Youth and Risk of Suicidal Ideation. Pediatrics. 2020;145(2):e20191725. https://doi.org/10.1542/peds.2019-1725.

52. Chew D, Anderson J, Williams K, May T, Pang K. Hormonal Treatment in Young People With Gender Dysphoria: A Systematic Review. Pediatrics. 2018;141(4):e20173742. https://doi.org/10.1542/peds.2017-3742.

53. de Vries Alc, McGuire JK, Steensma TD, Wagenaar ECF, Doreleijers TAH, Cohen-Kettenis PT. Young adult psychological outcome after puberty suppression and gender reassignment. Pediatrics. 2014;134(4):696–704. https://doi.org/10.1542/peds.2013-2958.

54. Amand CSt, Fitzgerald KM, Pardo ST, Babcock J. The Effects of Hormonal Gender Affirmation Treatment on Mental Health in Female-to-Male Transsexuals. J Gay Lesbian Ment Health. 2011;15(3):281–299. https://doi.org/10.1080/19359705.2011.581195.

55. Human Rights Campaign. Healthcare Equality Index 2020.; 2020.

56. Centers for Disease Control and Prevention. HIV in the Southern United States. 2019;29(September):1–4.

57. National Cancer Institute. Colorectal Cancer Screening: Cancer Trends Progress Report. Published April 2022. Accessed July 13, 2022. https://progressreport.cancer.gov/detection/colorectal_cancer.

58. Alsan M, Garrick O, Graziani G. Does Diversity Matter for Health? Experimental Evidence from Oakland. Am Econ Rev. 2019;109(12):4071–4111. https://doi.org/10.1257/aer.20181446.

59. Agénor M, Krieger N, Austin SB, Haneuse S, Gottlieb BR. At the intersection of sexual orientation, race/ethnicity, and cervical cancer screening: assessing Pap test use disparities by sex of sexual partners among black, Latina, and white U.S. women. Soc Sci Med. 2014;116:110–118. https://doi.org/10.1016/j.socscimed.2014.06.039.

60. Grasso C, Goldhammer H, Thompson J, Keuroghlian AS. Optimizing gender-affirming medical care through anatomical inventories, clinical decision support, and population health management in electronic health record systems. J Am Med Inform Assoc. 2021;28(11):2531–2535. https://doi.org/10.1093/jamia/ocab080.

61. Vermund SH, Wilson CM. Barriers to HIV testing--where next? Lancet. 2002;360(9341):1186–1187. https://doi.org/10.1016/S0140-6736(02)11291-8.

62. Rayment M, Thornton A, Mandalia S, et al. HIV Testing in Non-Traditional Settings - The HINTS Study: A Multi-Centre Observational Study of Feasibility and Acceptability. PLoS One. 2012;7(6):e39530. https://doi.org/10.1371/journal.pone.0039530.

63. Wee CC, McCarthy EP, Phillips RS. Factors associated with colon cancer screening: the role of patient factors and physician counseling. Prev Med. 2005;41(1):23–29. https://doi.org/10.1016/j.ypmed.2004.11.004.

64. Kirkoen B, Berstad P, Botteri E, et al. Acceptability of two colorectal cancer screening tests: pain as a key determinant in sigmoidoscopy. Endoscopy. 2017;49(11):1075–1086. https://doi.org/10.1055/s-0043-117400.

65. Kruk M, Matsick JL. A taxonomy of identity safety cues based on gender and race: From a promising past to an intersectional and translational future. Transl Issues Psychol Sci. 2021;7(4):487–510. https://doi.org/10.1037/tps0000304.

66. Gonzales G, Zinone R. Cancer diagnoses among lesbian, gay, and bisexual adults: results from the 2013-2016 National Health Interview Survey. Cancer Causes Control. 2018;29(9):845–854. https://doi.org/10.1007/s10552-018-1060-x.

67. MetLife. Still out, Still Aging: The MetLife Study of Lesbian, Gay, Bisexual, and Transgender Baby Boomers.; 2010.

68. American Medical Association. Creating an LGBTQ-friendly practice. Published 2021. https://www.ama-assn.org/delivering-care/population-care/creating-lgbtq-friendly-practice.

69. Cahill S, Makadon H. Sexual Orientation and Gender Identity Data Collection in Clinical Settings and in Electronic Health Records: A Key to Ending LGBT Health Disparities. LGBT Health. 2014;1(1):34–41. https://doi.org/10.1089/lgbt.2013.0001.

70. Gay & Lesbian Medical Association. Guidelines for Care of Lesbian, Gay, Bisexual, and Transgender Patients. Gay & Lesbian Medical Association Accessed July 15, 2022. https://www.glma.org/_data/n_0001/resources/live/Welcoming%20Environment.pdf.

71. Wilkerson JM, Rybicki S, Barber CA, Smolenski DJ. Creating a Culturally Competent Clinical Environment for LGBT Patients. J Gay Lesbian Soc Serv. 2011;23(3):376–394. https://doi.org/10.1080/10538720.2011.589254.

72. McClain Z, Hawkins LA, Yehia BR. Creating Welcoming Spaces for Lesbian, Gay, Bisexual, and Transgender (LGBT) Patients: An Evaluation of the Health Care Environment. J Homosex. 2016;63(3):387–393. https://doi.org/10.1080/00918369.2016.1124694.

73. Pratt-Chapman ML, Eckstrand K, Robinson A, et al. Developing Standards for Cultural Competency Training for Health Care Providers to Care for Lesbian, Gay, Bisexual, Transgender, Queer, Intersex, and Asexual Persons: Consensus Recommendations from a National Panel. LGBT Health. 2022;9(5):340–347. https://doi.org/10.1089/lgbt.2021.0464.

74. Sutter ME, Simmons VN, Sutton SK, et al. Oncologists’ experiences caring for LGBTQ patients with cancer: Qualitative analysis of items on a national survey. Patient Educ Couns. 2021;104(4):871–876. https://doi.org/10.1016/j.pec.2020.09.022.

75. Obedin-Maliver J, Goldsmith ES, Stewart L, et al. Lesbian, Gay, Bisexual, and Transgender-Related Content in Undergraduate Medical Education. JAMA. 2011;306(9):971–977. https://doi.org/10.1001/jama.2011.1255.

76. Keuroghlian AS, Ard KL, Makadon HJ. Advancing health equity for lesbian, gay, bisexual and transgender (LGBT) people through sexual health education and LGBT-affirming health care environments. Sex Health. 2017;14(1):119–122. https://doi.org/10.1071/SH16145.

77. Nowaskie DZ, Sewell DD. Assessing the LGBT cultural competency of dementia care providers. Alzheimers Dement N Y N. 2021;7(1):e12137. https://doi.org/10.1002/trc2.12137.

78. Cornelius JB, Carrick J. A Survey of Nursing Students’ Knowledge of and Attitudes Toward LGBT Health Care Concerns. Nurs Educ Perspect. 2015;36(3):176–178. https://doi.org/10.5480/13-1223.

79. Moll J, Krieger P, Moreno-Walton L, et al. The Prevalence of Lesbian, Gay, Bisexual, and Transgender Health Education and Training in Emergency Medicine Residency Programs: What Do We Know? Acad Emerg Med. 2014;21(5):608–611. https://doi.org/10.1111/acem.12368.

80. Greenwood BN, Carnahan S, Huang L. Patient-physician gender concordance and increased mortality among female heart attack patients. Proc Natl Acad Sci U S A. 2018;115(34):8569–8574. https://doi.org/10.1073/pnas.1800097115.

81. Laveist TA, Nuru-Jeter A. Is doctor-patient race concordance associated with greater satisfaction with care? J Health Soc Behav. 2002;43(3):296–306. https://doi.org/10.2307/3090205.

82. Wallis CJD, Jerath A, Coburn N, et al. Association of Surgeon-Patient Sex Concordance With Postoperative Outcomes. JAMA Surg. 2022;157(2):146–156. https://doi.org/10.1001/jamasurg.2021.6339.

83. Romanelli M, Hudson KD. Individual and systemic barriers to health care: Perspectives of lesbian, gay, bisexual, and transgender adults. Am J Orthopsychiatry. 2017;87(6):714–728. https://doi.org/10.1037/ort0000306.

84. Gonzales G, McKay T, Carpenter CS. Disparities in Health Insurance Coverage and Access to Care for Children By Mother’s Sexual Orientation. Matern Child Health J. 2020;24(5). https://doi.org/10.1007/s10995-019-02857-7.

